# Dynamics of SARS-CoV-2 VOC neutralization and novel mAb reveal protection against Omicron

**DOI:** 10.1101/2022.08.12.22278720

**Authors:** Linhui Hao, Tien-Ying Hsiang, Ronit R. Dalmat, Renee Ireton, Jennifer Morton, Caleb Stokes, Jason Netland, Malika Hale, Chris Thouvenel, Anna Wald, Nicholas M Franko, Kristen Huden, Helen Chu, Alex Greninger, Sasha Tilles, Lynn K. Barrett, Wesley C. Van Voorhis, Jennifer Munt, Trevor Scobey, Ralph S. Baric, David Rawlings, Marion Pepper, Paul K. Drain, Michael Gale

## Abstract

To evaluate SARS-CoV-2 variants we isolated SARS-CoV-2 temporally during the pandemic starting with first appearance of virus in the Western hemisphere near Seattle, WA, USA, and isolated each known major variant class, revealing the dynamics of emergence and complete take-over of all new cases by current Omicron variants. We assessed virus neutralization in a first-ever full comparison across variants and evaluated a novel monoclonal antibody (Mab). We found that convalescence greater than 5-months provides little-to-no protection against SARS-CoV-2 variants, vaccination enhances immunity against variants with the exception of Omicron BA.1, and paired testing of vaccine sera against ancestral virus compared to Omicron BA.1 shows that 3-dose vaccine regimen provides over 50-fold enhanced protection against Omicron BA.1 compared to a 2-dose regimen. We also reveal a novel Mab that effectively neutralizes Omicron BA.1 and BA.2 variants over clinically-approved Mabs. Our observations underscore the need for continued vaccination efforts, with innovation for vaccine and Mab improvement, for protection against variants of SARS-CoV-2.

**Summary:** We isolated SARS-CoV-2 temporally starting with emergence of virus in the Western hemisphere. Neutralization analyses across all variant lineages show that vaccine-boost regimen provides protection against Omicron BA.1. We reveal a Mab that protects against Omicron BA.1 and BA.2 variants.

## Introduction

SARS-COV-2 was first identified in the Western hemisphere near Seattle, WA, USA, with isolation of the North American ancestral virus, SARS-CoV-2/WA-1 in January 2020. Since then, the virus has undergone rapid genetic diversification leading to the progressive outgrowth of new virus strains(Harvey et al., 2021). Specific strains linked with epidemic coronavirus infectious disease 2019 (COVID-19), defined as variants of concern (VOCs) have progressively emerged, with the SARS-CoV-2 Omicron BA.1 variant first reported in November 2021. Compared to ancestral SARS-CoV-2/Wuhan and SARS-CoV-2/WA-1, the Omicron VOC harbors multiple mutations in the viral genome (Wolter et al., 2022), and Omicron infections have quickly spread globally accompanied by emergence of new subvariants (Araf et al., 2022; Wolter et al., 2022). Overall, SARS-CoV-2 has shown remarkable genomic diversification, with >1,500 unique Phylogenetic Assignment of Named Global Outbreak Lineages (PANGOLIN) (Rambaut et al., 2020). Some variants have demonstrated evidence of increased transmissibility, virulence, and/or immune evasion (Maeda et al., 1988), and the World Health Organization has labeled the Omicron variants among the latest VOCs. The large number of viral mutations for Omicron, many occurring in the spike protein, are linked with higher transmissibility and perhaps lower virulence than prior SARS-CoV-2 VOCs (Araf et al., 2022; Shuai et al., 2022). The spike protein is a mediator of host cell entry and the primary target of neutralizing antibodies (Harvey et al., 2021), Beta and Gamma SARS-CoV-2 variants have mutations that modestly reduce vaccine-induced neutralization (Garcia-Beltran et al., 2021). Similarly, Omicron variants have exhibited significantly reduced vaccine-induced neutralization as compared to ancestral SARS-CoV-2 (Wilhelm et al., 2021), and have enhanced affinity for angiotensin-converting enzyme 2 (ACE2), a virus co-receptor protein on target cells (Cele et al., 2021).

The three vaccines approved by the FDA for use in the United States all target the spike protein of the original ancestral SARS-CoV-2 first identified in Wuhan, China, which is conserved in the North American ancestral SARS-CoV-2/WA-1 strain (Feikin et al., 2022). These vaccines have been successful at inducing initial humoral immunity but have short lived neutralizing antibody response durability. The neutralizing antibody responses and vaccine effectiveness gradually decline after vaccination and may be negatively impacted by emerging VOCs (Bajema et al., 2021; Bar-On et al., 2021; Cromer et al., 2022; Khoury et al., 2021; Lopez Bernal et al., 2021; Tregoning et al., 2021). However, mRNA-based vaccine boosters can enhance neutralizing immunity (Bar-On et al., 2021; Doria-Rose et al., 2021; Garcia-Beltran et al., 2021), with clinical data supporting a level of vaccine effectiveness against Omicron BA.1 (Collie et al., 2022; Tseng et al., 2022). While the viral dynamics of VOC infections have been similar between vaccinated and unvaccinated persons (Kissler et al., 2021), the effectiveness of immunity from previous infection to prevent reinfection thus far has been substantially lower for Omicron (56%) as compared to the alpha (90%), beta (86%), and delta (92%) variants, indicating differences in immune evasion across these VOCs (Altarawneh et al., 2022). Indeed, a study of household SARS-CoV-2 transmission showed higher transmission for Omicron than Delta for fully vaccinated adults (Lyngse et al., 2021). While one study has shown the Omicron variant to have high neutralizing antibody escape (Schmidt et al., 2022), few studies have compared the neutralization activity between highly mutated variants, such as Omicron, and other VOCs among both vaccinated and unvaccinated adults.

We previously developed and validated a live virus neutralization assay to quantify differences in immunity from vaccine and host responses against SARS-CoV-2 (Rathe et al., 2021; Rodda et al., 2021). Here, we used our validated live virus neutralization assay to test sera from persons who had received one of three approved vaccines in the United States—mRNA-1273, BNT162b2, or Ad26.COV2.S—or had been infected with Omicron, another VOC, or ancestral SARS-CoV-2 form Seattle, WA, USA. We also directly compared the ability of antibody from mRNA vaccine two-dose recipients to neutralize representative virus of each major VOC, showing that while antibody neutralization is variable, it is effective across all VOCs with the exception of Omicron BA.1. We then show that convalescent sera do not effectively protect against Omicron BA.1, but vaccination following infection and recovery gives strong neutralization, and mRNA vaccination booster enhances neutralization of Omicron BA.1. We also define the properties of a novel human monoclonal antibody (Mab) that effectively neutralizes the Omicron BA.1 and Omicron BA.2 variants.

## Results

We established a clinic to isolate pipeline to characterize SARS-COV-2 variants directly from nasal swab samples collected at the primary point of care for diagnostic testing. We recovered and propagated SARS-CoV-2 primary isolates from the greater Seattle area representing the major VOCs. Figure 1A shows a phylogenetic analysis of a representative isolate of each Seattle VOC (O’Toole et al., 2021) in relation to ancestral SARS-CoV-2/Wuhan-Hu-1 and SARS-CoV-2/WA-1 strains. To evaluate mRNA vaccine-induced humoral immune efficacy to generally neutralize infection across SARS-CoV-2 variants, we first compared the ability of pooled sera from recipients of either mRNA-1273 (N=8) or BNT162b2 (N=7) 2-dose vaccination regimen, median 8 days prior, against immune sera from convalescent donors having recovered from previous SARS-CoV-2 infection, median 347 days since symptom onset (N=13) (Table 1), in a plaque reduction neutralization titer (PRNT) analysis using live virus of each VOC, prior to the emergence of Omicron BA.1. All VOCs tested were effectively neutralized by each vaccination sera pool. Paired T-test showed that no statistically significant difference between the Moderna and Pfizer mRNA-vaccines in virus neutralization. In contrast, pooled samples of sera collected from convalescent donors without prior vaccination showed significantly lower neutralization activities compared to either RNA vaccine (p<0.05) across all variants (Figure 1B).

**Figure 1.**
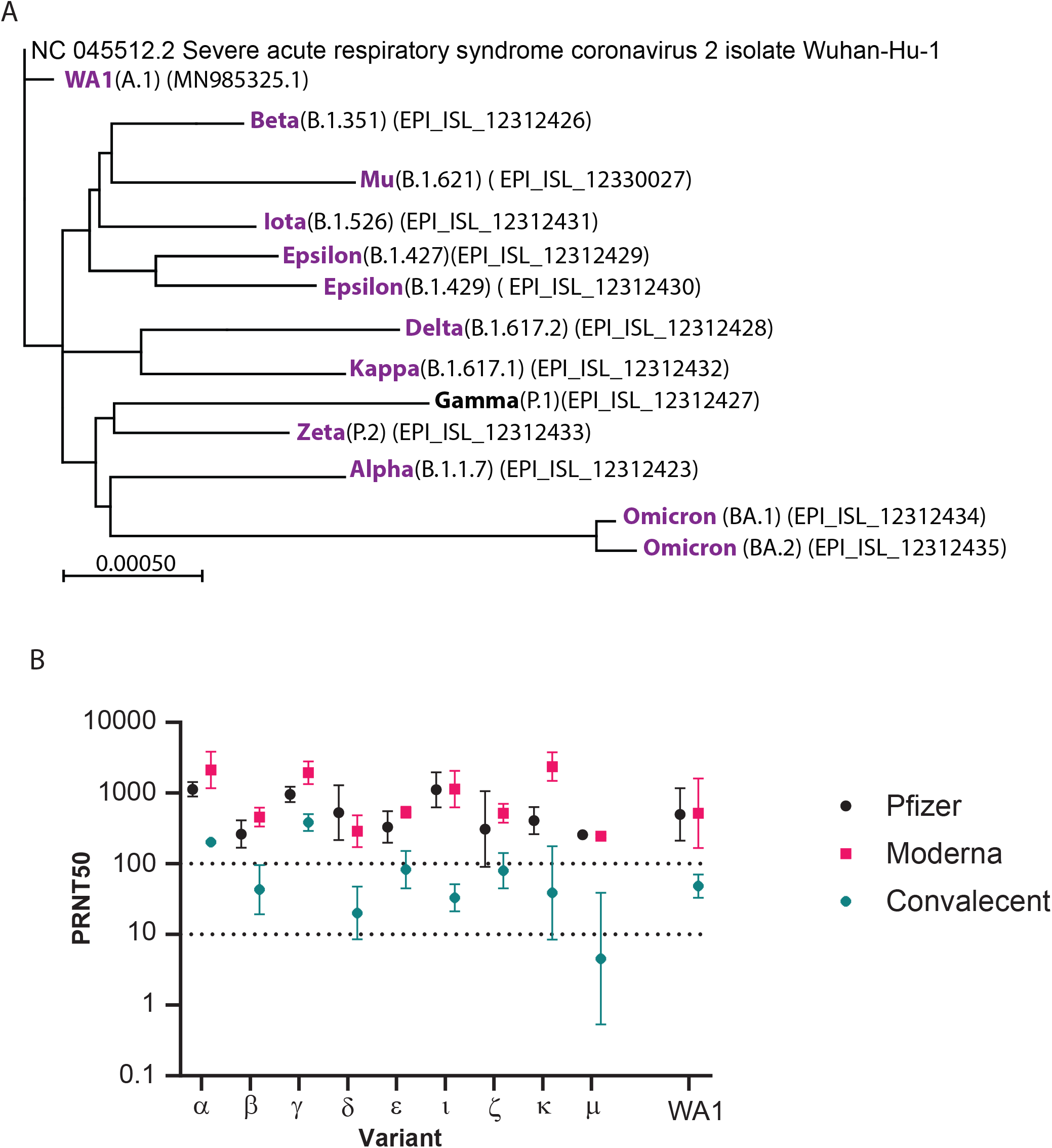
Sequence and neutralization comparison of SARS-CoV-2 variants. **A**. Phylogenetic tree showing relationships among SARS-CoV-2 variants isolated from patient samples in Seattle, WA, with the SARS-CoV-2/Wuhan-Hu-1 as a reference root. The tree is generated using online version of MAFFT(Katoh et al., 2017; Kuraku et al., 2013) and MEGAX(Kumar et al., 2018). Tree length is calculated with Maximum Pasimony method, the length of the branch is propotional to the number of substitutions. Scale denotes relative genetic distance. **B**. PRNT_50_ titer of pooled sera from SARS-CoV-2-naïve individuals vaccinated with Pfizer (BNT162b2) or Moderna (mRNA-12730) vaccine, and patients recovered from prior SARS-CoV-2 infection (convalescent; Table 1). Sera pools were tested for their neutralization activity against the indicated SARS-CoV-2 variants. Dotted lines show the 10 and 100 PRNT_50_ reference levels, with PRNT_50_ of 10 serving as the lower limit cutoff for virus neutralization activity.

**Table 1.** Characteristics of pooled sera sets from vaccinated or convalescent serum donors.

|  | Vaccinated (two-dose) | Convalescent |
| --- | --- | --- |
| Characteristics | N=15 | N=13 |
| Sex |  |  |
| Male | 7 (46.7%) | 2 (15.4%) |
| Female | 8 (53.3%) | 11 (84.6%) |
| Age |  |  |
| Min-Max | 25-77 | 22-62 |
| Median (IQR) | 35.5 (30-47.8) | 45.5 (38-53) |
| Vaccine type |  |  |
| mRNA-1273 | 8 (53.3%) |  |
| BNT162b2 | 7 (46.7%) |  |
| None |  | 13 (100%) |
| Time since | Recent vaccine dose | Symptom onset |
| Min-Max | 6-20 days | 157 - 366 days |
| Median (IQR) | 8 (8-12.5) days | 347 (289.5 – 360.5) days |

In the beginning of December 2021, we first detected the presence of the Omicron variant in Seattle using the S-gene target failure (SGTF) PCR assay (Figure 2A), following the tail of the Delta variant infections (Pangolin BA.1.617.2). We isolated and sequenced the Omicron variant from these SGTF-identified early cases. By mid-December 2021, the Omicron variant became predominant (Figure 2A, lower). While the Seattle Omicron variant is highly similar to the Omicron VOC BA.1 first reported from South Africa (Viana et al., 2022), the Seattle Omicron additionally encodes an S1265I nsp3 substitution and a three amino acid deletion (aa 105-107) in nsp6. We also identified three novel sites of amino acid deletion in the spike-coding region (aa 68-69, aa 142-144, and aa 211) and an R346K substitution. Moreover, the Nucleocapsid protein (NP) encoded a novel deletion of aa 30-47 (Figure 2B). 3-D modeling of the BA.1 spike protein and spike protein binding to *ACE2* placed these aa changes in context of spike protein structure (Figure 2C).

**Figure 2.**
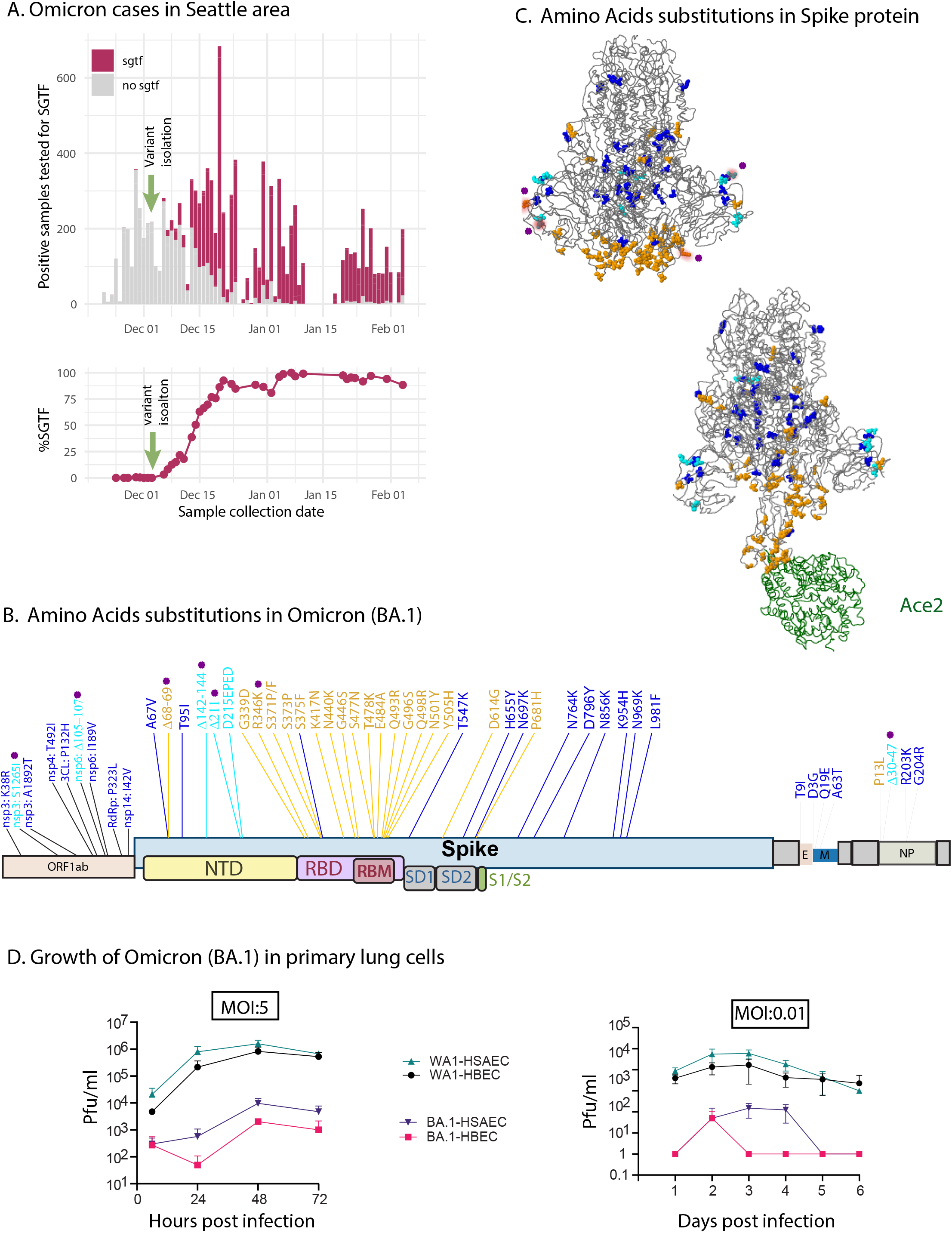
Sequence and characterization of Seattle Omicron strain. **A**. S gene target failure RT-PCR assay (SGTF; TaqPath assay(Perchetti et al., 2021)was applied for new variant surveillance. Upper: Histogram shows the positive cases of Omicron strain emerging in Seattle starting in December 2021 into February 2022. Lower: Isolation of the Omicron BA.1variant was isolated and confirmed by viral genome sequencing, replacing other circulating variants for new cases by early January 2022. **B**. Aa sequence changes in the Seattle Omicron BA.1 variant compared to the ancestral Wuhan-Hu-1 and WA-1 SARS-CoV-2 strains. Purple denotes novel mutations specific for this Seattle Omicron BA.1 variant. Aa changes occurring in more than 100 known Omicron BA.1 sequences appear in blue colored font. If the aa change occurs at a site known to be involved in phenotypic effects such as altering host-cell receptor binding or antigenicity, it is shown as orange. Aa insertion or deletions are colored in cyan. C. 3D model of Omicron BA.1 spike protein was generated using analysis tool on GISAIS website(Khare et al., 2021). Aa color follows the same rule as in B. **D**. Single step (left) and multi-step (right) growth and production of infectious virus by SARS-CoV-2/WA-1 and Seattle Omicron BA.1 variant in human lung bronchial epithelial cells (HBEC) and human lung small airway epithelial cells (HSEC). Plaque titers were measured from culture supernatant over the time course shown. Paired t-test was performed, showing that BA.1 produces significantly lower amount of virus compared to ancestral strain at 12-48 hours post infection

Infection analysis in primary human bronchial epithelial cells (HBEC) and human primary small airway epithelial cells (HSAEC) revealed the growth properties of the Omicron BA.1 variant compared to ancestral SARS-CoV-2/WA-1. One-step virus growth analysis demonstrated markedly slower replication and significantly lower peak virus production by Omicron BA.1 (Figure 2D left). Multi-step virus growth analysis revealed slower virus replication and spread by Omicron BA.1 (Figure 2D right) but exhibited overall better growth in HSAEC compared to HBEC. These results indicate that the Omicron BA.1 VOC has significantly reduced replication fitness in lung epithelial cells compared to ancestral SARS-CoV-2.

To determine the sensitivity of the Omicron BA.1 VOC compared with ancestral virus to humoral immunity from SARS-CoV-2 infection and recovery prior to the emergence of Omicron BA.1 (convalescence) or to convalescence followed by vaccination, we evaluated sera from convalescent persons or convalescent persons who received the 2-dose vaccine regimen after their recovery from SARS-CoV-2 infection. For this study convalescence was defined as median 157 days since symptom onset for all persons (Table 2). Using PRNT_50_ titer of 10 as a lower end limit for virus neutralization (Gilbert et al., 2022), we conducted paired PRNT analysis to evaluate neutralization antibody titer against Omicron BA.1 and ancestral virus. We used the neutralization titer mean for each virus to calculate neutralization differential (ND) value representing the fold increase of neutralization PRNT_50_ for ancestral virus compared to Omicron BA.1. Across individual sera samples, we found that convalescent sera provided only low PRNT_50_ titer (range 1-100) with antibodies that were less effective at neutralizing Omicron BA.1 and exhibited variable ability to neutralize SARS-CoV-2/WA-1 (Figure 3A). However, vaccination with either mRNA-1274, BNT162b2, or JNJ-78436735 following convalescence typically provided an increase in PRNT levels that demonstrated enhanced protection against both Omicron BA.1 and ancestral SARS-CoV-2/WA-1. In non-vaccinated convalescent persons, the mean PRNT_50_ level was below or at the lower limit for neutralization of either virus, with a nonsignificant trend for enhanced neutralization of ancestral virus compared to Omicron BA.1. However, in convalescent persons who received any of the three major vaccines the ND value ranged from 2.8 to 3.5, with significant differences in PRNT_50_ titer between Omicron BA.1 and ancestral virus (Figure 3B). As an additional comparator, we included evaluation of four persons who received a 2-dose prime/boost inactivated virus vaccine regimen of Sinovac (Akova and Unal, 2021) or SinoPharm (Xia et al., 2021) COVID-19 vaccine, with serum collected median 165 days post-second vaccine dose (Table 2B) (Palacios et al., 2020). Samples obtained from persons who received inactivated virus did not have detectable PRNT antibody titer.

**Table 2.**
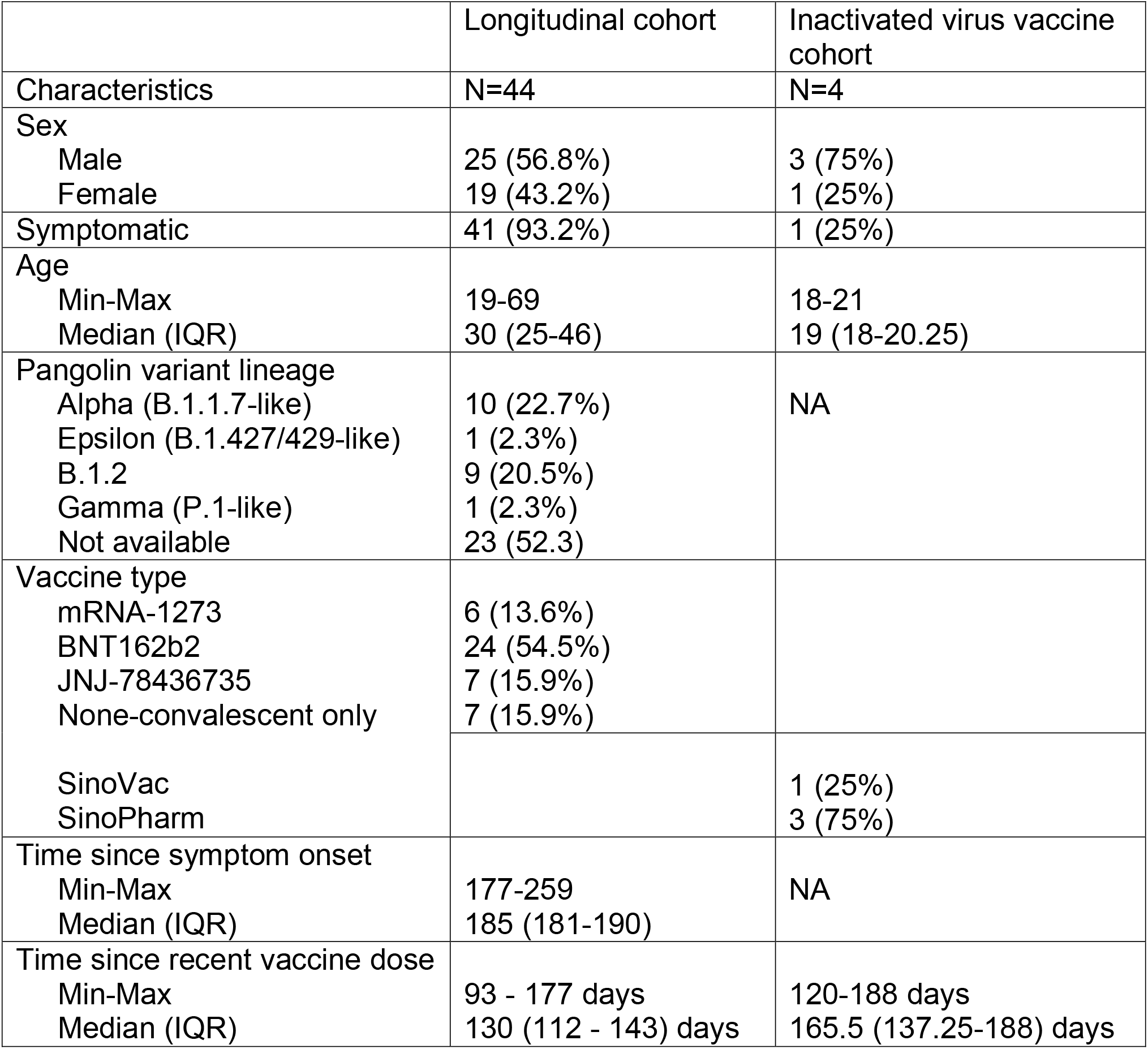
Characteristics of convalescent-vaccination serum donors.

**Figure 3.**
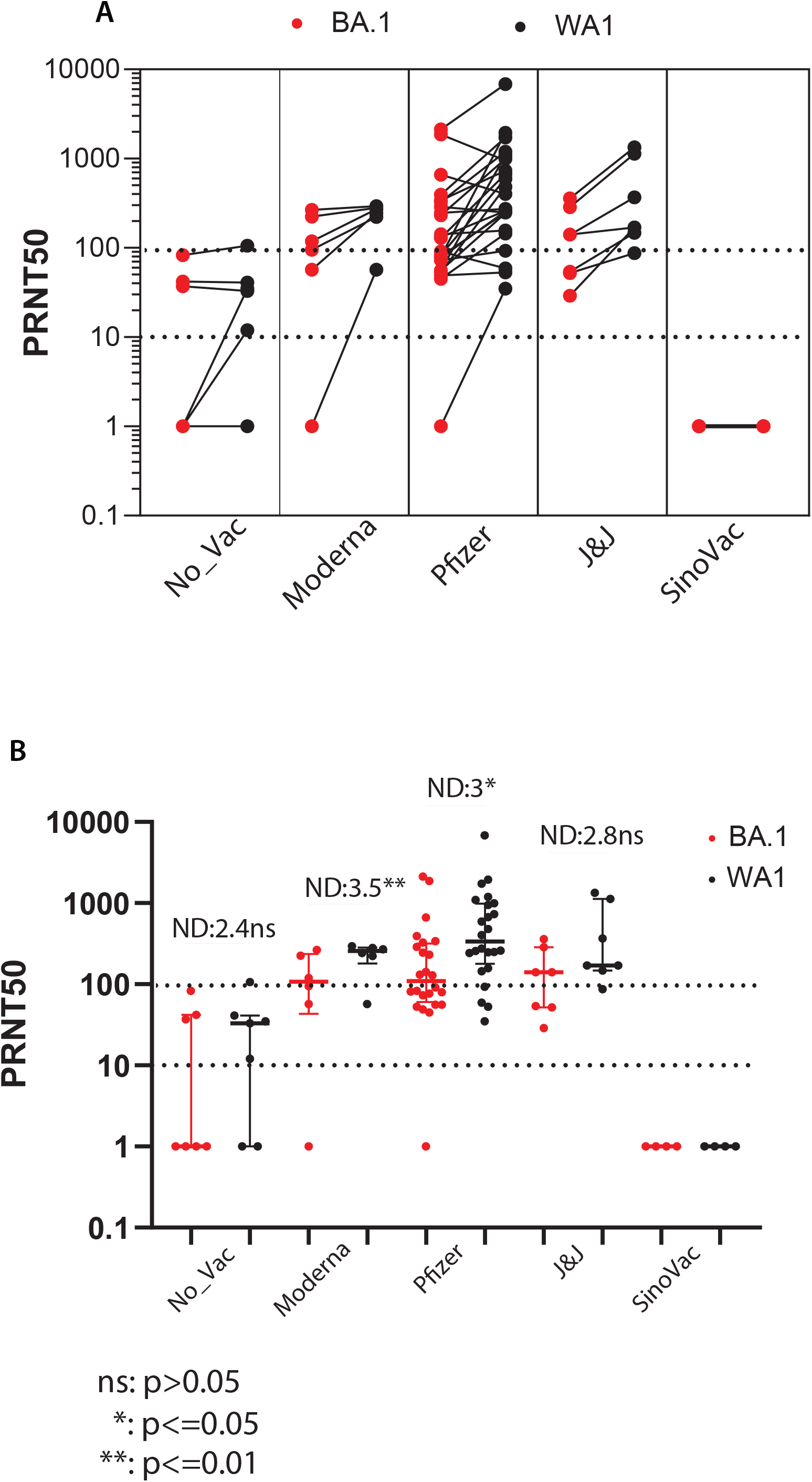
Convalescent sera do not protect against Omicron but vaccination following infection and recovery enhances humoral immunity for virus neutralization. Serum samples were provided from a longitudinal cohort of SARS-CoV-2 infection. Convalescent individuals either received no vaccine or were vaccinated with the indicated vaccines (Table 2). Sera were evaluated for virus neutralization by PRNT_50_ assay against SARS-CoV-2/WA-1 and the Seattle Omicron BA.1 variant in side-by-side paired testing. **A**. Mean PRNT_50_ titers of individual sera in paired testing against each virus. **B**. PRNT_50_ data of serum analyses showing mean and all input data points. Neutralization difference (ND) values were calculated by dividing mean PRNT_50_ value of SARS-CoV-2/WA-1 neutralization by the PRNT_50_ mean of Omicron BA.1 neutralization for each sera set. PRNT_50_ values were statistically evaluated using paired T-test. P values are shown for each comparison. The PRNT_50_ 10 lower limit neutralization value and 100 value are marked by the dotted lines.

We evaluated the ability of two and three-dose vaccination regimens to induce neutralizing antibody responses against the Omicron BA.1 VOC in SARS-CoV-2-naïve persons. We conducted paired PRNT_50_ analyses of sera collected from vaccine recipient cohorts who had no previous SARS-CoV-2 infection but received two dose vaccination, median 153 days previously or from three dose vaccination of each vaccine with sera collected at median 32 days since the third vaccine dose (Table 3). Importantly, this timeline links closely to the emergence of the Omicron BA.1 variant and the approval for use of the vaccine booster. We generated side-by-side PRNT_50_ values for each individual for neutralization of Omicron BA.1 and ancestral SARS-CoV-2/WA-1 infection. As shown in the Figure 4A, neutralizing antibody PRNT_50_ values (including values near the neutralization cutoff) in persons receiving the 2-dose vaccine regimen were consistently lower against Omicron BA.1 compared to ancestral virus. In contrast, sera from vaccination/boost (3-dose) regimen exhibited an increase in PRNT_50_ against both Omicron BA.1 and ancestral virus. Evaluation of mean PRNT_50_ levels revealed an ND of 20 with 2-dose vaccination. In contrast, vaccination with 3-dose regimen demonstrated enhanced neutralization of Omicron BA.1 to nearly completely overlapping levels with ancestral virus neutralization. Remarkably, the ND value of this comparison was reduced to 3.5 compared to ND 20 for 2-dose vaccine regimen (Figure 4B). Comparison of 3-dose to 2-dose vaccine regimen for each virus alone revealed major enhancement of Omicron BA.1 neutralization (ND=54) over enhancement of ancestral virus neutralization (ND=9.4) and demonstrated that neutralization of each isolate was significantly improved by the 3-dose vaccination.

**Table 3.** Characteristics of donors received two-dose regimen or three-dose regimen.

|  | Two-dose | Three-dose |
| --- | --- | --- |
| Characteristics | N=6 | N=10 |
| Sex |  |  |
| Male | 2 (33.3%) | 2 (20%) |
| Female | 4 (66.7%) | 8 (80%) |
| Age |  |  |
| Min-Max | 31-61 | 22-63 |
| Median (IQR) | 50 (37-56.75) | 29 (24-36.75) |
| Vaccine type |  |  |
| BNT162b2, BNT162b2 | 6 (100%) |  |
| mRNA-1273, mRNA-1273, mRNA-1273 |  | 8 (80%) |
| BNT162b2, BNT162b2, mRNA-1273 |  | 2 (20%) |
| Time since recent vaccine dose |  |  |
| Min-Max | 152 - 161 days | 19-50 days |
| Median (IQR) | 153.5 (153-157) | 32 (27-34.5) days |

**Figure 4.**
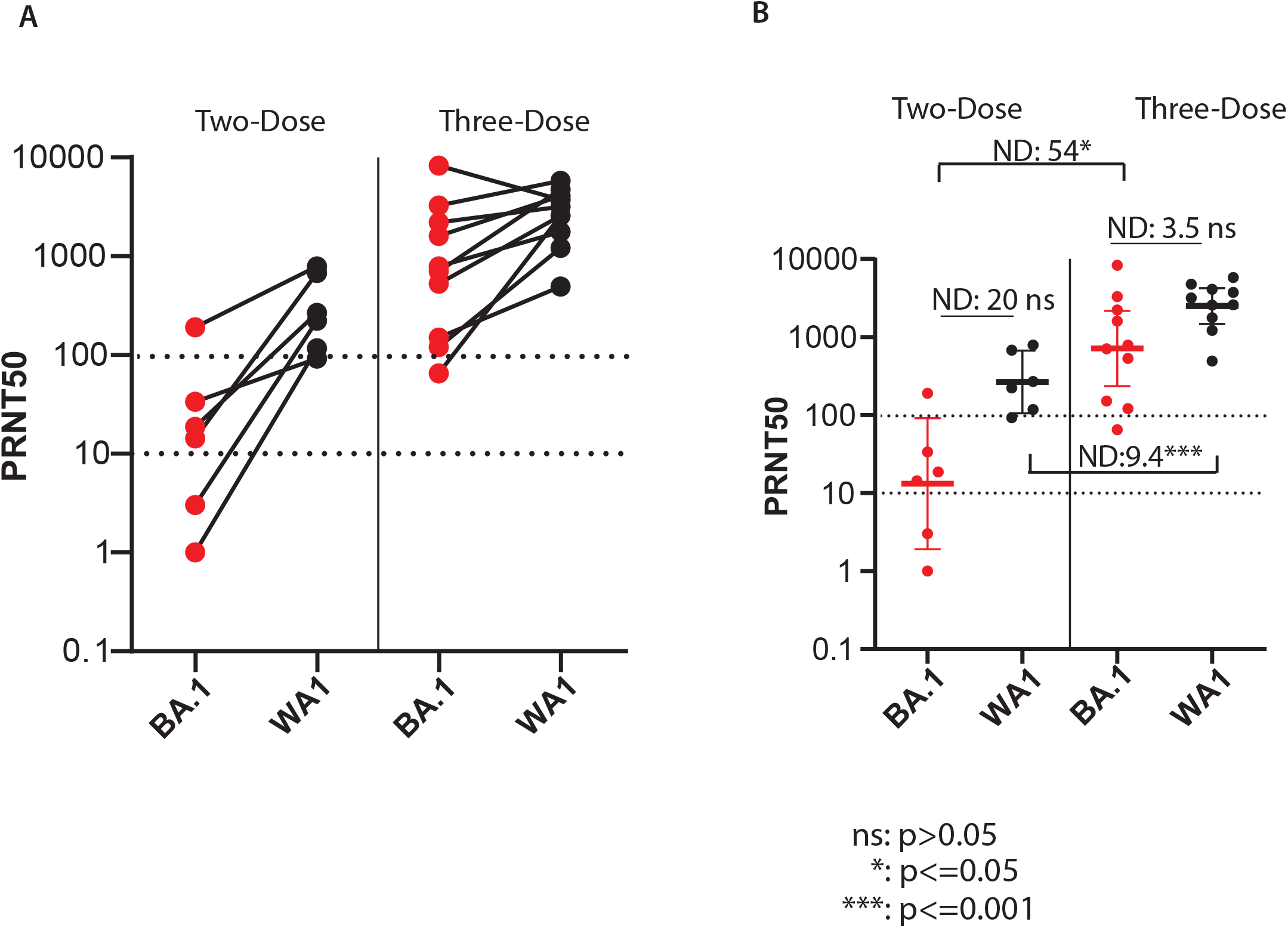
Vaccination/boost regimen enhances protection against Omicron. PRTN_50_ titers were determined from serum samples from SARS-CoV-2 naïve individuals who received vaccination 2-dose regimen (Pfizer vaccine) or 3-dose vaccine regimen (Moderna or Pfizer with Moderna boost; Table 3). Neutralization of SARS-CoV-2/WA-1 and the Seattle Omicron BA.1 variant were determined in by side paired testing. **A**. Mean PRNT_50_ titers of individual sera in paired testing against each virus. **B**. PRNT_50_ data of serum analyses showing mean and all input data points. Neutralization difference (ND) values are shown. PRNT_50_ values were statistically evaluated using paired T-test. Significant differences are indicated with p value. Ns, nonsignificant. The PRNT_50_ 10 lower limit neutralization value and 100 value are marked by the dotted lines. ns: p>0.05, *: p<=0.05, ***: p<=0.001

As humoral immunity is currently the hallmark for protection against SARS-CoV-2 infection, we and others have developed human monoclonal antibodies to leverage as immune-therapeutics to treat infection (Hunt et al., 2022; VanBlargan et al., 2021; Zhou et al., 2022). We recently described human monoclonal antibodies (Mabs) against SARS-CoV-2 that were isolated via a cloning strategy to recover and produce antibodies from convalescent persons infected with ancestral SARS-CoV-2/WA-1 in the first wave of the USA epidemic in Seattle (Rodda et al., 2021). To identify possible Mabs that can neutralize Omicron, we screened a panel of Mabs for neutralization activity against SARS-CoV-2/WA-1, Delta, and Omicron BA.1. During the preparation of this manuscript, the Omicron variant BA.2 emerged in Seattle, and was isolated and sequenced for inclusion in the Mab neutralization assessment. Compared to the first wave Seattle Omicron BA.1 variant, the emergent Omicron BA.2 variant displays additional aa substitutions. As shown in Figure 5A, 6 new amino acids substitutions appeared in the Omicron BA.2 spike protein alone. 20 amino acids in the spike protein are different between the Omicron BA.1 and Omicron BA.2 Seattle isolates. We evaluated the sensitivity of these Omicron variants to Mab neutralization. As controls, we compared the Mabs against the clinically-used Regeneron REGN10987/10933 two mAB cocktail (Weinreich et al., 2021) and a negative control antibody (MaliA01) against a *Plasmodium falciparum* protein, the latter used to establish the non-neutralizing threshold (Thouvenel et al., 2021). To determine neutralization activity, we conducted focus reduction neutralization titer (FRNT) analysis to identify the dose of each antibody that conferred 50% reduction of SARS-CoV-2 infection foci in A549 cells expressing ACE2 and TEMPRSS2 (FRNT_50_), with the SARS-CoV-2 Delta variant as a comparator control. Within this panel, we identified four Mabs that neutralized ancestral virus and the Delta variant. Remarkably, one of these Mabs (Mab 297) had neutralization activity against both Omicron BA.1 and Omicron BA.2 variants. (Figure 5A). The Regeneron REGN10987/10933 mAB cocktail had no neutralizing activity against the Omicron variants. We also tested our Mabs for neutralization activity against SARS-CoV (Wec et al., 2020) and bat coronavirus SHC014, with direct comparison to SARS-CoV-2, each from infectious clone virus encoding a nano-luciferase reporter, to determine Mab inhibitory dose (ID_50_) (Menachery et al., 2015). As an additional control, we included the ADG2 Mab that is known to neutralize all three viruses (Rappazzo et al., 2021). With the exception of the MaliA01 control, all the test Mabs neutralize the ancestral SARS-CoV-2 but not SARS-CoV. Collectively, these results show that Mab297 can neutralize across Omicron BA.1 and Omicron BA.2 variatns, Delta, and ancestral virus to a level that is comparable to the neutralization activity of Regeneron REGN10987/10933 mAB cocktail against ancestral virus only. While REGN10987/10933 does not neutralize the Omicron BA.1 nor Omicron BA.2 virus, Mab297 does neutralize both VOCs. Moreover, Mab297 had increased potency to neutralize ancestral and Delta variants at a FRNT_50_ dose less than 10 ng/ml.

**Figure 5.**
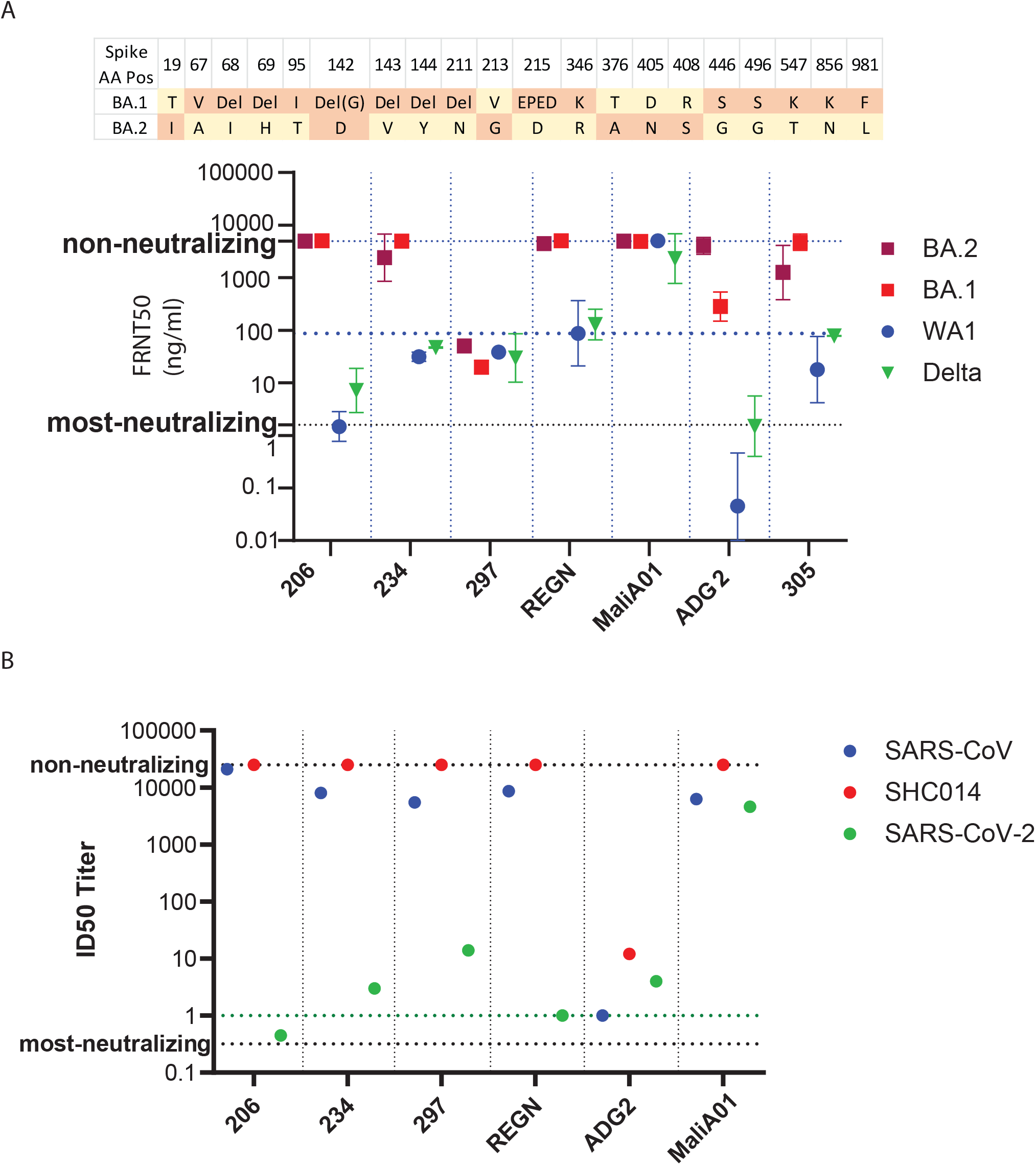
Mab neutralization. **A. Upper**: Spike protein aa changes comparing Omicron BA.1 and Omicron BA.2 variants. Aa position (AA Pos) in the Spike protein is indicated by position number. Del, deletion. Site variation and insertion sequence are shown for aa positions 142 and 215, respectively. **Lower**: FRNT_50_ in ng/ml Mab concentration shown for the indicated Mabs. REGN, Regeneron 10987/10933 mAB cocktail. MaliA01 is a negative control antibody against a *Plasmodium falciparum* protein and is used to establish the non-neutralizing threshold. ADG2 is a positive-control Mab. Lower, middle, and upper dotted lines respectively mark the FRNT_50_ levels for most neutralizing, neutralizing, and non-neutralizing concentration, respectively, of the test Mabs. **B**. ID_50_ titer analysis of virus neutralization. Lower, middle (green) and upper dotted lines respectively mark the ID_50_ titer level for most-neutralizing, ADG2 neutralization of SARS-CoV (positive control), and non-neutralizing thresholds.

## Discussion

In this study, we isolated every major class of SARS-CoV-2 variant present in Seattle, including all known classes and VOCs except lambda, which has not been known to circulate in the Seattle area. We report the emergence of the Omicron BA.1 variant and its epidemic takeover of SARS-CoV-2 cases from the previously dominating Delta variant, followed by emergence of Omicron BA.2 variant. The Seattle Omicron BA.1 variant shares the sequence of the B.1.1.529 original virus but this variant has two new aa substitutions and five sites of single or multiple aa deletions, including a 17 aa deletion in the NP coding region. In the spike protein, we identified an R346K substitution in the receptor binding domain and 5 aa deletions (see Figure 2B). These changes in the Omicron BA.1 spike protein likely contribute to the lack of antibody neutralization demonstrated by this variant against sera from convalescent and vaccinated person as it became the predominate strain in early 2022 of the Seattle SARS-CoV-2 epidemic.

At the time Omicron BA.1 cases were surging in Seattle there were about three quarter of a million cumulative SARS-CoV-2 cases in Washington (WA) state, more than 50% of people in WA has been fully vaccinated, and certain group of people had the third dose booster. To assess the population immunity against the emergence of the Omicron BA.1 variant, we compared antibody neutralization of Omicron BA.1 and ancestral SARS-CoV-2/WA-1 in paired testing, showing that sera from convalescent individuals have a reduced ability to neutralize SARS-COV-2 compared to vaccinated individuals, wherein neutralization activity against Omicron BA.1 is low or nonexistent. Though our cohort size was small, we show that sera from convalescent individuals who later received Moderna, Pfizer or J&J vaccination possess variable neutralization activity against Omicron BA.1 and have an ND approximately 3 or greater for neutralization of ancestral virus. Our data clearly show that among SARS-CoV-2 infection-naïve persons who received the 2-dose vaccine regimen, the ability to neutralize Omicron BA.1 variant was approximately 20-fold lower compared to neutralization activity against ancestral virus. Importantly, these data show that receiving one vaccine boost clearly closes the gap and reduces the ND between Omicron BA.1 and ancestral virus, in addition to improving protective antibody titers against both viruses. In fact, we found that the ND values from the Omicron BA.1 PRNT_50_ assay were increased by approximately 54-fold when comparing sera from 2-dose versus 3-dose vaccine regimen. These observations demonstrate that vaccine boost delivers a significant and functional enhancement in protective antibody levels against both Omicron BA.1 and ancestral virus infection in boosted persons.

We identified that a previously described human Mab, Mab297 (Hale et al., 2022; Rodda et al., 2021), effectively neutralizes Omicron variants BA.1 and Omicron BA.2, the Delta variant, and the ancestral SARS-CoV-2/WA-1. In our assays, the single antibody performed better than the Regeneron REGN10987/10933 mAB cocktail comprised of two different Mabs that do not neutralize these Omicron variants. The ability of Mab297 to neutralize Omicron BA.1 and Omicron BA.2 variants underscores a potential clinical application for this Mab in treatment of COVID patients to neutralize specific lineages of the Omicron variants including BA.1 and BA.2 viruses and protect against disease. Despite the additional changes in the Seattle Omicron BA.1 and Omicron BA.2 variants, Mab297 possesses neutralizing activity suggesting it binds to epitope(s) conserved across ancestral, Delta and Omicron variants. Further comparative studies that assessed neutralization of SARS-CoV and SHC014 against bat-derived virus show that Mab297 is not effective against other SARS coronavirus. In contrast, the positive control the ADG2 Mab neutralized SARS-CoV-2 ancestral virus and delta variant, SARS-CoV, and SHC014 CoV but not SARS-CoV-2 Omicron variants, indicating that the ADG2 Mab recognizes an evolutionary conserved epitope on the SARS-CoV-2 RBD overlapping with the hACE2 binding, and suggesting that this epitope is altered in the Omicron variants (Iketani et al., 2022; Rappazzo et al., 2020).

Taken together, our study reveals that the SARS CoV-2 Omicron variant produces blunted neutralizing antibody responses compared to ancestral strains and suggests a mechanism by which Omicron evades host immune responses. Therapeutic strategies such as additional vaccine boosters or monoclonal antibodies targeting conserved regions in the viral proteome may be effective measures to overcome immunological escape elicited by SARS CoV-2 variants.

## Materials and methods

### Isolation of SARS-CoV-2 variants from clinical samples

SARS-CoV-2 nasal swab specimens were collected by the University of Washington Virology Laboratory and saved in viral transport media (VTM)or PBS. Virus-positive samples with diagnostic RT-qPCR cycle threshold (Ct) < 33 were identified from reference testing on the Roche cobas, Hologic Panther Fusion, and Abbott Alinity-m platforms(Lieberman et al., 2020). 400µL of PBS/VTM from SARS-CoV-2 positive specimens were extracted using the MagMax Viral/Pathogen Kit on the KingFisher Flex (Thermo) and RT-qPCR was performed using the TaqPath COVID-19 Combo Kit on the ABI 7500, following manufacturer’s instructions. Positive specimens were transferred to UW biosafety level (BSL)3 laboratory for virus culture. For virus isolation, the VTM was filtered through Corning Costar Spin-X centrifuge tube filter (CLS8160), and 0.1 ml of the filtered VTM was used to infect VeroE6 cells ectopically expressing human Ace2 and TMPRSS2 (VeroE6-AT cells; a gift from Dr. Barney Graham, National Institutes of Health, Bethesda MD) in a 48-well plate. A typical cytopathic effect related to SARS-Cov2 infection was normally observed within 2-4 days post infection. Virus-containing culture supernatants were collected and designated as a passage P0 virus stock. P1 virus stock cultures were grown in VeroE6/TMPRSS2 cells (JCRB1819) using P0 virus as seeding inoculum. The titer of the P1 stock was measured by standard SARS-CoV-2 plaque assay as described (Rathe et al., 2021)(Addetia et al., 2020).

### Sequencing

P1 stock virus was verified with whole genome sequencing analysis. An aliquot of P1 stock was subject to RNA extraction (Zymo Research, R1040) and used as template to produce cDNA with SuperScript™ IV First-Strand Synthesis System (ThermoFisher, Waltham, MA, USA). The products were then used in library production using the Swift SARS-CoV148 2 SNAP Version 2.0 kit (Swift Biosciences™, Ann Arbor, MI, USA) following the manufacturer’s instructions. Resulting libraries were quality-assessed using the Agilent 4200 TapeStation (Agilent Technologies, Santa155 Clara, CA, USA). Libraries with concentrations of 1.0 ng/µL were sequenced on an Illumina NextSeq 500 (Illumina, San Diego, CA, USA) along with positive and negative controls. The resulting sequence data were processed through covid-swift-pipeline (https://github.com/greninger-lab/covid_swift_pipeline). Consensus genome sequence were generated by aligning the amplicon reads to the SARS-CoV-2 Wuhan-Hu-1 ancestral reference genome (NC_045512.2). For each genome, at least 1 million raw reads were acquired, representing >750x mean genome coverage and a minimum of 10x base coverage. Each consensus genome was then analyzed to assign lineage based on the Pangolin dynamic lineage nomenclature scheme (O’ Toole et al., 2021).

### Virus growth characterization

The growth kinetics of SARS-CoV-2 variants were observed in human lung bronchus epithelial cells HBEC3-KT (ATCC, CRL-4051) ectopically expressing human *ACE2* and human lung small airway epithelial cells HSAEC1-KT (ATCC, CRL-4050) ectopically expressing human *ACE2*. In brief, 5×10^5^ cells were seeded in 24-well plates the day before infection. On the day of experiment, the plates were transferred to the UW BSL3 facility, and the cells were first rinsed once with DPBS, and then inoculated with designated SARS-CoV-2 virus variants in 250ul basal medium (PromoCell C-21060, C-21070) at multiplicity if infection (MOI) of 5 for one-step growth curves, and MOI 0.01 for multistep growth curves. After virus adsorption at 37°C for 1 hour, the inoculum was removed and complete growth media (basal medium with supplements) for the cells was then added to the cells. For one-step growth curve analysis, the culture medium was collected at 6, 24, 48 and 72 hours post infection, and virus replication was quantitated using plaque assay. The virus production for multi-step growth curve was measured on 1 to 6 days post infection in quadruplets replicates. Growth curves were generated using GraphPad Prism 9.3.1.

### Monoclonal antibody isolation and characterization

We recently reported that spike protein receptor binding domain (RBD) memory B cells were enriched in convalescent blood donors three months following SARS-CoV-2 infection, from which we isolated single RBD-specific B cells and sequenced the B cell receptors from three persons. We cloned paired heavy and light chain sequences that were obtained, and expressed them as IgG1 monoclonal antibodies (Rodda et al., 2021). Antibodies were expressed in small scale cultures and assessed for antibody expression and RBD binding by enzyme-linked immunosorbent assay (ELISA) specificity by RBD ELISA. RBD-binding antibodies were further assessed for their capacity to inhibit RBD binding to the ACE2 receptor by surrogate Virus Neutralization Test assay (Rodda et al., 2022; Rodda et al., 2021). We selected monoclonal antibody (Mab)297 for further analyses based on best performance in these screening assays. Mab297 and control Mabs were expressed on a larger scale and purified as described (Rodda et al., 2021) for more detailed analyses.

### Antibody Neutralization Assay

Whole blood samples were collected from participants of multiple study cohorts spanning the duration of the SARS-CoV-2 pandemic from 4/2020-12/2021 (see Table 1 for details on sera donors). Individuals with and without prior history of SARS-CoV-2 infection were followed longitudinally throughout vaccination with mRNA-1273, BNT162b2 and JNJ-78436735, SinaVac and SinaPharm. Samples were collected 6 months after primary vaccination series and 15 days after administration of a third dose. The serum was obtained from whole blood samples by removing the red blood cells through centrifugation. All serum samples were heat inactivated by incubating at 56°C for 1 hour. The protection ability of serum and Mabs against SARS-CoV-2 was assessed by Plaque Reduction Neutralization Test (PRNT) or Focus-forming-unit Reduction Neutralization Test. Briefly, serial 5-fold dilution of each antibody containing sample was prepared with starting dilution of 1:5 for serum or appropriate concentration for monoclonal antibody into gelatin saline (0.3% [w/v] gelatin in phosphate buffered saline supplemented with CaCl_2_ and MgCl_2_). Each antibody dilution was mixed with equal volume of gelatin saline containing 60 pfu virus for PRNT, or 100 pfu for FRNT. The antibody-virus mixture was incubated at 37°C for 1 hour and then used in a plaque assay with VeroE6/TMPRSS2 cells as described (Addetia et al., 2020; Rathe et al., 2021) to calculate PRNT_50_.

For FRNT, A549-hACE2-TMPRSS2 (Invivogen a549-hace2tpsa) cells were plated in 96-well plates at 0.1 ml /well at 0.15×10^6^ cells/ml in Dulbecco’s Modified Eagle Media (DMEM) supplemented with 10%FBS, 1X Penicillin/Streptomycin/(Pen/Strep), 0.1 mg/ml Normocin (Invivogen), 0.5 µg/ml puromycin (Invivogen), and 0.1 mg/ml hygromycin (Invivogen) the day before performing the assay. The seeding medium was removed, and the antibody-virus mixture was added to each well. The plate was incubated for 1 hour at 37°C to allow virus attachment, and then an equal volume of media composed of DMEM with 4% heat-inactivated Fetal Bovine Serum (FBS) and 1% Penicillin/Streptomycin/L-Glutamate was added. The cells were incubated at 37°C for 20 hours prior to fixation with 100% methanol for 15 minutes at room temperature. The fixative was removed, and the plate was blocked with 50 µl/well blocking buffer (PBS containing10% FBS,5% non-fat dry milk, and 0.1%Tween-20) for 60 minutes at room temperature. Anti-SARS-CoV-2 N (MA5-36086) diluted 1:10000 in primary antibody buffer (0.1 % Saponin, 0.1% BSA, 0.05% Tween-20 in PBS) was used to detect focus-forming units by adding 30 µl of antibody buffer to each well and incubating at 4°C overnight. The primary antibody was washed off and the secondary antibody, HRP conjugated goat anti-rabbit IgG (ImmunoReagents, GtxRb-003-FHRPX), at 1:10,000 dilution in PBS was added for incubation at 4°C overnight. The focus-forming units were visualized using TrueBlue™ Peroxidase substrate (SeraCare, 5510-0030) and imaged and counted with Cytation V (Agilent, CA). FRNT50 was calculated using GraphPad Prism 9.3.1 by employing non-linear regression analysis.

For antibody neutralization of SARS-CoV-2 encoding Nano-luc reporter gene (SARS-CoV-2-Nluc), antibodies were first diluted serially in a 96-well plate using a Beckman I-5 automation system. The plate was then transferred to the BSL-3 laboratory for infection with 800 PFU/mL SARS-CoV-2-Nluc virus per well. The plate containing antibodies and virus was mixed and incubated for 1 hour at 37°C. The antibody-virus mixture was then added to a 96-well plate with each well pre-seeded with Vero cells and incubated for 18 hours at 37°C. Reduction in relative light units (RLU) given by the nanoluciferase reporter was measured using the Promega Nano-Glow Luciferase Assay System, with output captured and recorded on a luminometer. The inhibitory dose (ID_50_) titer for each antibody was determined using a custom Excel macro to process data set output, and data were graphically processed and visualized with GraphPad Prism, V8 using non-linear regression analysis.

### Ethics Statement

This study was conducted with approval by the institutional review board at the University of Washington. Participants received standard-of-care measures according to local and institutional guidelines. Some participants were enrolled in the Hospitalized or Ambulatory Adults with Respiratory Viral Infections (HAARVI) study (STUDY00000959) and/or the Healthy Adult Specimen Repository study (STUDY00002929), the Point of Care study (STUDY00009981), or the Protocol for the Collection of Laboratory Research Specimen (STUDY00004312). Blood collection for the purpose of neutralizing antibody analyses was also conducted under approval number STUDY000098108. Subjects were recruited via print advertisements, provided with information about the study, risks associated, and how their privacy would be protected. To enroll in the study each subject or their legally-authorized representative provided verbal understanding and written consent.

## Data Availability

All data produced in the present work are contained in the manuscript

## Acknowledgements

Supported by NIH grant AI100625 (MG) and AI151698 (MG & WCVV, UWARN).

## Author Contributions

Conceptualization: L. Hao, M.J. Gale, P.K. Drain, M. Pepper, and R.S. Baric. Investigation and data analysis: L. Hao, T. Hsiang, J. Munt, and T. Scobey. Clinical data and specimens acquistition: R. Dalmat, J. Morton, C. Stokes, A. Greninger, A. Wald, N.M. FranKo, K. Huden, H. Chu, P.K. Drain, S. Tilles, L.K. Barrett, W.C. Van Voorhis, J. Netland, M. Hale, C. Thouvenel, D. Rawlings, M. Pepper. R. Ireton provided critical research infrastructure. L. Hao, M.J. Gale, P.K. Drain prepared and edited the manuscript. All authors provided critical feedback and approval of the final manuscript.

